# Association of RDoC dimensions with post-mortem brain transcriptional profiles in Alzheimer’s Disease

**DOI:** 10.1101/2024.10.07.24315057

**Authors:** Weiqian Jiang, Jonathan Vogelgsang, Shu Dan, Peter Durning, Thomas H. McCoy, Sabina Berretta, Torsten Klengel

**Author notes:** Correspondence: Sabina Berretta MD, McLean Hospital, 115 Mill Street, Belmont MA 02478 and Torsten Klengel MD, PhD, McLean Hospital, 115 Mill Street, Belmont MA 02478. Denotes equal contribution (Weiqian Jiang, Jonathan Vogelgsang, Sabina Berretta, Torsten Klengel). **Author contributions:** WJ, JSV, TK and SB designed the study. WJ, JSV, SD, and PD performed analyses. WJ, JSV, TK and SB wrote the manuscript. All authors approved the final version of the manuscript.

## Abstract

**INTRODUCTION:** Neuropsychiatric symptoms are common in people with Alzheimer’s disease (AD) across all severity stages. Their heterogeneous presentation and variable temporal association with cognitive decline suggest shared and distinct biological mechanisms. We hypothesized that specific patterns of gene expression associate with distinct NIMH Research Domain Criteria (RDoC) domains in AD.

**METHODS:** Post-mortem bulk RNAseq on the insula and anterior cingulate cortex from 60 brain donors representing the spectrum of canonical AD neuropathology combined with natural language processing approaches based on the RDoC Clinical Domains.

**RESULTS:** Distinct sets of >100 genes (p_FDR_<0.05) were specifically associated with at least one clinical domain (Cognitive, Social, Negative, Positive, Arousal). In addition, dysregulation of immune response pathways was shared across domains and brain regions.

**DISCUSSION:** Our findings provide evidence for distinct transcriptional profiles associated with RDoC domains suggesting that each dimension is characterized by specific sets of genes providing insight into the underlying mechanisms.

## Introduction

Alzheimer’s disease (AD) is a progressive neurodegenerative disorder resulting in dementia. While advanced stages of AD are characterized by severe cognitive impairment, varying degrees of neuropsychiatric symptoms (NPS) including depression, agitation, aggression, and apathy can be present across the entire spectrum of AD^1,2^. NPS can lead to accelerated disease progression, increased caregiver burden, and earlier death. Treatment of these symptoms with medications used for non-neurodegenerative psychiatric disorders is based on the largely unchallenged assumption that their biological underpinnings are equivalent. The clinical diagnosis of AD is based on a combination of clinical and pathological measurements, either biomarkers in cerebrospinal fluid (CSF) or PET imaging of Amyloid-β or TAU deposition. Neuropathologically, AD is characterized by extracellular plaques of misfolded β-amyloid (Aβ) aggregates and intracellular neurofibrillary tangles (NFT) formed by paired helical filaments of hyperphosphorylated tau protein^3^. While deposition of Aβ and NFT are considered neuropathological hallmarks of AD^4^, studies on transcriptional changes in AD using post-mortem brain tissue suggest the dysregulation of multiple cellular pathways including synaptic dysfunction, gliosis, demyelination, and inflammation leading to neuronal loss^5-9^. Bridging between canonical and molecular changes on one side and symptoms on the other is particularly challenging, even more so in the context of clinical, genetic, and neuropathological heterogeneity among persons with AD^10^.

Most large-scale transcriptional studies focus on extensively studied brain regions such as hippocampus, anterior cingulate cortex (BA32/BA33) and prefrontal cortex (BA9, BA10, BA46)^6-9,11,12^. In contrast, the insula, which is located deep within the lateral sulcus that separates the temporal from the parietal and frontal lobes, remains largely understudied. It has been traditionally viewed as a paralimbic or limbic integration cortex integrating visceral information^13,14^. Recently, imaging studies have sparked considerable interest in investigating the role of the insular cortex in the context of emotion, pain, decision making, motor control, and social functions^15^. The insular cortex is connected to a wide variety of brain regions including the frontal, anterior cingulate and parietal cortex, limbic areas such as the amygdala, hypothalamus, and entorhinal cortex, and to sensorimotor brain regions^16,17^. The central role of the insular cortex in relevant circuits is supported by several neuroimaging studies suggesting its involvement in AD and AD-related NPS^18-21^. However, transcriptomic studies investigating molecular mechanisms underlying pathological alterations of the insular cortex in AD remain scarce.

To date, most transcriptomic studies focus on the comparison between normative individuals and people with either early- or late-onset AD^5,8,12^. Single-nucleus RNAseq studies have provided significant insight into AD pathology by revealing distinct gene expression patterns in multiple cell types^22,23^. Although very informative, categorical diagnosis comparison designs may not be well suited to account for interindividual heterogeneity in symptom presentation, particularly regarding comorbid NPS. To overcome these limitations, we previously applied a natural language processing (NLP) algorithm to post-mortem medical records of donors with and without AD to provide dimensional phenotyping within the context of the NIMH Research Domain Criteria (RDoC)^24^. RDoC is a multidimensional framework composed of different neuropsychiatric domains capturing a spectrum of symptoms rooted in brain circuits and biology^25-27^.

In this study, we performed an RNAseq analysis in the anterior Insula (aINS; BA16) and dorsal anterior cingulate cortex (dACG; BA32), focusing on a dimensional approach based on the NIMH RDoC domain matrix including Negative Valence Systems, Positive Valence Systems, Cognitive Systems, Social Processes, and Arousal and Regulatory Systems. Using a generalized linear regression model, we show that transcriptomic changes associated with dimensional RDoC clinical domain scores provide a deeper and more nuanced insight into the underlying molecular correlates of dimensional symptoms presentation in AD. Importantly, our results suggest common and distinct molecular mechanisms across RDoC domains and brain regions.

## Methods

### Experimental Subjects and Tissue Preparation

All tissue samples and medical records were obtained from the Harvard Brain Tissue Resource Center (HBTRC; operating under the MGB McLean Institutional Review Board (IRB)). The subject cohort (n=60) available for this study included brain donors representing the full spectrum of Braak & Braak neuropathological stages, thus representing the neuropathological AD progression from Braak and Braak stages 0 to II (unaffected controls) to Braak and Braak stages III and IV (mild to moderate AD pathology) and Braak and Braak stages V and VI (severe AD pathology)^28^. Only donors with sufficient medical records and a life diagnosis by a qualified clinician were included. Donors with significant psychiatric conditions diagnosed during adolescence, early- or mid-adulthood were not included. Similarly, we excluded donors with non-AD neurological diagnoses.

Medical records in hard copies were scanned into computer-readable text files using optical character recognition. Text files were processed using RDoC-based NLP algorithms to obtain quantitative measures of clinical domain scores, as described by McCoy et al^24^ (*https://github.com/thmccoy/CQH-Dimensional-Phenotyper*). Access to donors’ medical records and other sensitive data was restricted to IRB-authorized investigators; all other investigators contributing to this study were given access to de-identified data, according to the Health Insurance Portability and Accountability Act (HIPAA) regulations. Detailed metadata information for each sample is given in **Supplementary Data 1**. A summary of the cohort subject basic data by RNA-seq batch is included in **Supplementary Data 2** and individual scores in each domain is given in **Supplementary Data 3**.

The dACG (BA32) and the aINS (BA16) were isolated from flash-frozen human post-mortem brain tissue samples. Tissue blocks were sectioned using a cryostat and 5 × 40μm sections (approximately 20 mg) were collected for RNA extraction using Absolutely RNA Miniprep Kit (Agilent, Lexington, MA) according to the manufacturer’s protocol. Briefly, 400μl lysis buffer, including 2.8μl β-mercaptoethanol were added to the tissue and homogenized. 400μl 70% ethanol was added to the tissue homogenate and vortexed for 5 seconds. 400μl homogenate was transferred to an RNA Binding Spin Cup and centrifuges for 60 seconds at 16,000 x g and repeated once. The column was washed once with 600μl Low-Salt Wash Buffer and centrifuged at max speed to dry, followed by a 15-minute DNase I digestion at room temperature. Next, the column was washed with 600μl High-Salt Wash Buffer, 600μl Low-Salt Wash Buffer, and 300μl Low-Salt Wash Buffer, each step followed by 60 seconds centrifugation at max. speed. Lastly, 40μl Elution Buffer was added on the matrix and incubated for 120 seconds followed by centrifugation at maximum speed for 60 seconds.

### RNA-Seq library preparation and sequencing

RNA library preparation and sequencing were conducted at Azenta Life Sciences (South Plainfield, NJ, USA). Briefly, RNA samples were quantified using Qubit 2.0 Fluorometer (ThermoFisher Scientific, Waltham, MA, USA) and RNA integrity was checked with 4200 TapeStation (Agilent Technologies, Palo Alto, CA, USA). RNA samples were treated with TURBO DNase (Thermo Fisher Scientific, Waltham, MA, USA) to remove DNA following manufacturer’s protocol. rRNA depletion sequencing libraries were prepared by using QIAGEN Fastselect HMR. RNA sequencing library preparation uses NEBNext Ultra II RNA Library Preparation Kit for Illumina by following the manufacturer’s recommendations (NEB, Ipswich, MA, USA). Briefly, enriched RNAs were fragmented for 15 minutes at 94 °C. First strand and second strand cDNA were subsequently synthesized. cDNA fragments were end repaired and adenylated at 3’ends, and universal adapters are ligated to cDNA fragments, followed by index addition and library enrichment with limited cycle PCR. Sequencing libraries were validated using the Agilent Tapestation 4200 (Agilent Technologies, Palo Alto, CA, USA), and quantified using Qubit 2.0 Fluorometer (ThermoFisher Scientific, Waltham, MA, USA) as well as by quantitative PCR (KAPA Biosystems, Wilmington, MA, USA). Sequencing libraries were multiplexed and clustered onto a flow-cell. After clustering, the flow-cell was loaded onto the Illumina Novaseq instrument according to manufacturer’s instructions. The samples were sequenced using a 2×150bp Paired End (PE) configuration. Image analysis and base calling were conducted by the Control Software. Raw sequence data (.bcl files) generated from Illumina was converted into fastq files and de-multiplexed using Illumina bcl2fastq 2.20 software. One mismatch was allowed for index sequence identification.

### Processing of RNA-seq data

RNA-seq data was processed and analyzed using the bcbio-nextgen Bulk RNA-seq pipeline (available at https://github.com/bcbio/bcbio-nextgen). Paired-end sequence reads were aligned to human genome UCSC GRCh38.p14 using STAR^29^. Quality metrics were assessed using Samtools^30^, with an average mapping rate of 84%. No samples were excluded based on sequencing QC metrics. Transcript counts from aligned reads were quantified using Salmon^31^, then summarized to the gene level using the R package Tximport^32^. Sequencing depth was normalized between samples using the median of ratios method in R package DESeq2^33^. Low expressed genes with less than 10 reads in over half of the total sample were removed.

### Differential Expression Analysis over Clinical Domain Scores

Scores for RDoC-based clinical domains were derived using NLP algorithms as previously described in Vogelgsang et al.^34^ The clinical domain scores of all donors were binned between 0 and 1, in 0.1 intervals and considered on a dimensional scale from 0 (lower limit) to 1 (upper limit) (**Supplementary Data 3**). Note that higher scores indicate more severe symptoms (e.g. more severe cognitive impairment); thus, a positive correlation between gene expression and clinical domain scores indicates that higher gene expression is associated with greater symptom severity. Differential gene expression analyses in the aINS and dACG for over dimensional scores for each of the 5 clinical domains using the filtered gene set was performed by ImpulseDE2^35^.

Briefly, after filtering, normalized counts were used in a principal component analysis (PCA) to identify potential covariates. The first five PCs were correlated with known technical and biological variables, resulting in significant correlations for neuropathological Braak & Braak stage, brain region, sex, age, RIN, and sequencing batch, but not PMI (**Supplementary Figure 1**). To further explore residual unknown sources of variation including the influence of changing cell type composition, medication, and comorbidities, surrogate variable analysis (SVA)^36^ was used, which yielded two significant SVs correlated with the first five PCs. Given the overall influence of Braak & Braak stages, brain regions, sex, age, RIN, sequencing batch, SV1, and SV2 on the data, we further explored the relevance of covariates across the RDoC cognition domain for each brain region and found that sex, sequencing batch and Braak & Braak status showed significant associations, while age, RIN, PMI, SV1, and SV2 were not significantly associated across the dimension (**Supplementary Data 1**). Thus, the final model to identify genes progressively regulated along the axis from low to high clinical domain scores in ImpulseDE2 included sex and sequencing batch as covariates. Braak and Braak stage was not included due to collinearity with the clinical domain scores. Statistical significance of differential expression was defined at p_FDR_<0.05.

**Figure 1.**
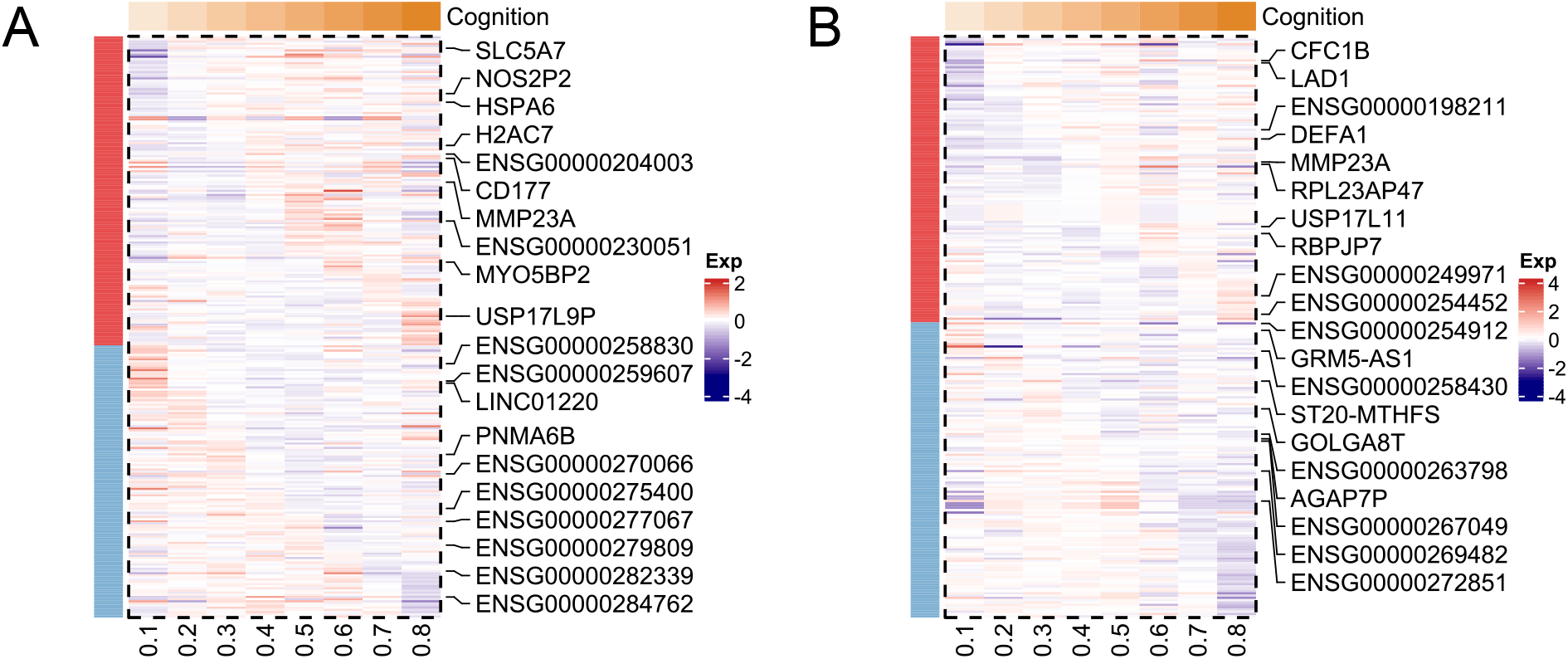
Transcriptional signature of dimensional RDoC cognition profiling. (**A**) Heatmap showing gene expression signatures of all DEGs (p_nominal_<0.05) with a significant up- or down-regulated pattern over increasing RDoC cognition values in the dACG. The top ten up- or down-regulated DEGs (all p_FDR_<0.05) are labeled. Bar colors on the left side indicate whether genes are clustered in an increasing (red) or decreasing (blue) trajectory. (**B**) Heatmap showing gene expression of DEGs (p_nominal_<0.05) with a significant up- or down-regulated pattern over increasing RDoC cognition values in the aINS. Top ten up- or down-regulated monotonous DEGs (all p_FDR_<0.05) are labeled.

### Functional Enrichment Analysis

A one-tailed hypergeometric test was conducted for pathway enrichment analysis using Metascape^37^. We included multiple databases such as Gene Ontology (GO), Kyoto Encyclopedia of Genes and Genomes (KEGG), Reactome and Wikipathway, separately for up- and downregulated differentially expressed (DE) genes from the dimensional analyses. Statistical significance was defined at p_FDR_<0.05.

### Data Availability

RNAseq data is available through the GEO accession number GSE261050. The code of the analyses is available at the Klengel Lab GitHub page under https://github.com/klengellab/RDoC_RNAseq.

## Results

### Differential Gene Expression in aINS and dACG as a Function of Dimensional Cognition Scores

In a previous proof-of-concept work, we used NLP algorithms to obtain cognition scores and showed that they are significantly associated with neuritic plaque load across all lobes of the brain^38^. The correlation of this classical neuropathological hallmark of AD with compelling evidence for a direct relationship to cognition^28,39,40^ and cognition scores provided evidence for the feasibility and validity of post-mortem dimensional phenotyping of brain donor electronic health records beyond categorical diagnoses, allowing for a more granular investigation of neuropsychiatric symptoms in AD.

Because the donor cohort available for this study only partially overlaps with the prior one^38^, we again tested the association between neuritic plaque load and cognitive symptom burden. As expected, the cohort analyzed here showed a significant association between neuritic plaque load across all lobes and cognition scores, as well as a significant association of Braak & Braak stages with cognition scores (**Supplementary Data 4**).

To investigate the underlying molecular changes associated with dimensional cognition burden, we regressed cognition scores over gene expression in the dACG and aINS of all donors, representing the full spectrum of Braak & Braak stages (from 0 to VI) using ImpulseDE2^35^. In the aINS, 109 DEGs at p_FDR_<0.05 showed a monotonous increase or decrease of expression across cognition score (**Figure 1 A, Supplementary Data 5**). Out of these 109 genes, 67 genes were positively correlated with more severe cognitive impairment. Top hits included genes involved in innate immune response, such as *CD177* and *HSPA6*. The remaining 42 genes were negatively correlated with cognition scores. These included *PNMA6B*, a member of paraneoplastic Ma antigen (PNMA) family, which is associated with immune-related diseases and neurological disorders^41,42^, alongside a large number of non-coding RNAs (ncRNAs) and pseudogenes.

In dACG, 107 DEGs (p_FDR_<0.05) showed a monotonous increased or decreased expression in association with cognition scores (**Figure 1 B, Supplementary Data 5**). Among the 48 genes with increased expression associated with more severe cognition scores, *LAD1* and *MMP23A* were the top hits, with a strong positive correlation with cognitive impairment. Similar to the aINS, top downregulated DEGs primarily belong to the less known group of pseudogenes or ncRNAs, with a total of 59 genes showing significant negative correlation with cognition severity.

### Differential Gene Expression in aINS and dACG as a Function of Clinical Domain Scores

Next, we focused on the Arousal Regulatory, Negative Valence, Positive Valence, and Social Systems domain scores. As expected, all domains were highly intercorrelated (Pearson r=0.88 ± 0.04). As shown in **Figure 2 A** and **Supplementary Data 5**, each clinical domain was associated with distinct sets of DEGs (all p_FDR_<0.05). In the aINS, DEGs were detected in association with Arousal (56 genes increased; 45 genes decreased), Negative valence (40 genes increased; 56 decreased), Positive valence (61 genes increased; 61 genes decreased) and Social domain (61 genes increased; 49 genes decreased). Similar number of DEGs were detected in dACG in association with Arousal (42 genes increased; 52 genes decreased), Negative valence (65 genes increased; 60 genes decreased), Positive valence (46 increased; 55 genes decreased) and Social domain (55 genes increased and 65 genes decreased (**Figure 2 B and Supplementary Data 5**). Notably, the majority of DEGs in aINS and dACG were uniquely associated with one domain with moderate to minimal overlap between domains (**Figure 2 C and D Supplementary Data 5 and 6**).

**Figure 2.**
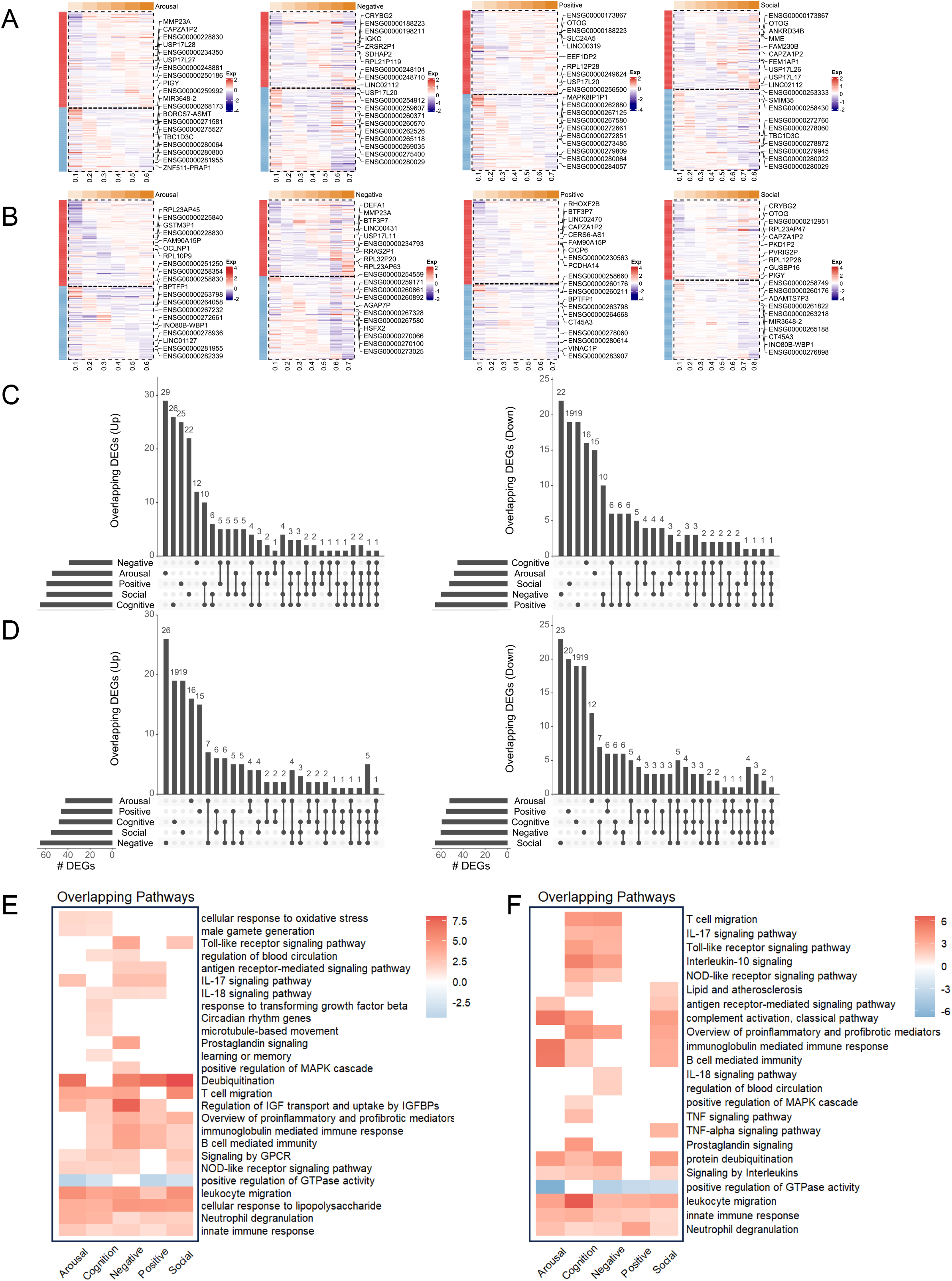
Identification of gene expression programs as a function of distinct RDoC cldomains. (**A**) Heatmaps showing expression trajectories of up- or down-regulated DEGs (p_nominal_<0.05) over increasing RDoC domain values separated for arousal, negative, positive and social in the dACG. Top ten up- or down-regulated DE genes (all p_FDR_<0.05) labeled. (**B**) Heatmaps showing analogous expression pattern in the aINS, separated for arousal, negative, positive and social. (**C**) Upset plots showing the overlap of DEGs of each RDoC domain, for increasing (left) and decreasing (right) genes, in the aINS. Overlapping DEGs are indicated by intersecting lines among different domains. (**D**) Upset plots showing overlapping DEGs for each RDoC domain, for increasing (left) and decreasing (right) genes, in the dACG. (**E**) Heatmap showing overlapping and distinct regulatory pathways for each RDoC domain in aINS and dACG (**F**). Colors indicate whether pathways are up- (red) or down-regulated (blue), and color depths represent significance levels of the enriched pathways.

### Pathway Enrichment in aINS and dACG as a Function of Clinical Domain Scores

Next, we assessed functional pathways across all clinical domains in aINS and dACG. Across both brain regions and all clinical domains, we found a predominating enrichment of pathways related to immune system functions. Enrichment analyses in the aINS revealed multiple pathways associated with innate immune responses shared across the five domains (**Figure 2 E, Supplementary Data 7**), including neutrophil degranulation, LPS response and leukocyte migration, supporting the notion that immune dysregulation is a driving factor for AD disease progression across all clinical domains investigated. In contrast to the broad association of innate immune response pathways with all clinical domains, other functional pathways were unique to a specific domain or a subset of domains. Specifically, pathways associated with the cognition domain included those involved in learning and memory, microtubule-based movement, circadian rhythm, and response to TGF-β. Moreover, pathways related to sex hormones were unique to the arousal, cognition, and negative domain. As an example, FSHB, beta subunit of follicle-stimulating hormone was specifically increased in domain arousal and cognition. This hormone, crucial in the female reproductive cycle as well as in stimulating production and maturation of sperm in males, has been shown to accelerate amyloid-β and Tau deposition in neurons and to impair cognition in an AD rodent model^43^, and sex steroid hormones have been associated with AD onset as well as progression^44^. Very similar shared biological processes were detected among the five domains in BA32 (**Figure 2 F, Supplementary Data 7**). Unique pathways included complement activation shared by domain arousal, cognition and social, which has been associated with Aβ clearance^45^.

## Discussion

Historically, investigations on AD placed emphasis on the pathogenesis of canonical findings such as neurofibrillary tangles and senile plaques. Although the relevance of β-amyloid and tau proteinopathies is uncontroversial, growing evidence supports the contribution of additional molecular pathways such as immune regulation, oxidative stress, insulin signaling and lipid metabolism, synaptic regulation and sex hormone signaling^46,47^.

The relationship between AD neuropathology and dysregulation of molecular pathways on the one hand and AD symptomatology on the other hand has been predominantly investigated in the context of categorical diagnosis frameworks and with strong emphasis on cognitive impairment. However, categorical diagnoses have been criticized for their limitations and their potential to impede scientific progress^48^. Over the last decade, efforts have been made to overcome these limitations by establishing dimensional models of human cognition, behavior and emotions that are based on neurobiological or behavioral phenotypes such as RDoC or HiTOP^49,50^. Although these efforts are not without controversy, dimensional phenotyping across diagnostic entities is a promising approach to identify molecular mechanisms behind distinct symptoms and syndromes. Distinct neuropathological and molecular patterns may, at least in part, account for the heterogenous clinical presentation of neuropsychiatric disorders including AD^51^.

We previously investigated the relationship between continuous cognitive symptom dimension in AD and classical neuropathological changes. Our results confirmed an association of NLP-derived cognitive scores with hallmark neuropathological findings and provided a proof-of-concept supporting the validity of NLP-based methodologies to obtain quantitative measures of functional RDoC domains from post-mortem health records. Here we link dimensional clinical domain scores to bulk RNAseq data on 101 post-mortem brain samples from aINS and dACG to test the hypothesis that gene expression signatures are associated with dimensional phenotype constructs derived from post-mortem brain donor health records. Our results provide evidence for gene expression signatures that define each clinical domain and brain region, potentially facilitating future research into more granular phenotypes beyond categorical diagnoses. Our data also suggest immune-related transcriptional changes as a common underlying mechanism across all domains and brain regions.

Results of linear regression models across all clinical domains in both brain regions yielded between n=94 and n=125 DEGs associated with one of the domains (**Supplemental Data 5**). Comparisons of up- or downregulated DEGs showed only a moderate level of overlap between clinical domains, suggesting that each domain may be defined by a specific set of differentially expressed genes (**Figure 2 C and D**). Notably, immune response pathways were robustly dysregulated across all clinical domains. These findings contribute to growing evidence for a critical role of immune signaling factors in AD and suggest their pervasive contribution to the overall clinical presentation of this disorder.

We detected a substantial number of FDR-significant DEGs with an increasing or decreasing expression pattern over cognition scores in aINS (n=109 DEGs) and dACG (n=107 DEGs) (**Figure 1 A and B**). In the aINS, a subset of genes was uniquely differentially expressed in association with the cognitive domain (upregulated n=26, downregulated n=16, **Figure 2 C, Supplementary Data 7**) including *H2AC7* and *LCN2* (both upregulated). *H2AC7*, a H2A histone protein variant, is involved in regulating cell cycle processes. Interestingly, reactivation of cell cycle related genes and DNA double-strand breaks are early pathological hallmarks of AD, and eventually lead to neuronal loss^52,53^. *LCN2* is involved in a wide range of biological processes such as regulation of iron homeostasis, inflammation, cell death, survival, differentiation and migration^54^. LCN2 in the brain has been implicated in cognition and behavior while increased levels of LCN2 are associated with age-related CNS diseases such as AD and PD^55^. In the dACG, a total of 38 genes were uniquely differentially expressed with the cognitive domain (upregulated n=19, downregulated n=19, **Figure 2 D, Supplementary Data 7**). Top hits include *LAD1* and *MMP23A* which showed a strong positive correlation with cognition scores. LAD1 is involved in cell anchoring and adhesion and MMP23A is one of the matrix metalloproteinase (MMP) family engaged in cell adhesion and matrix degradation in the extracellular matrix (ECM), enhancing immune cells migration and inflammatory response^56^.

DEGs that are uniquely up- or down-regulated over specific clinical domains may shed light on underlying molecular mechanisms of NPS in AD. For example, *PINX1* and *HSPA7 showed* a unique signature of increased expression over the arousal domain in the aINS. *PINX1* can inhibit the activity of telomerase, which is protective against ROS production and oxidative stress at different stages of AD pathology^57^. Interestingly, telomerase activity is also associated with circadian oscillation under the control of CLOCK-BMAL1 heterodimers^58^, a molecular mechanism fundamental to the arousal domain. *HSPA7* is a member of the human Hsp70 family and overexpression of Hsp70 can have protective effects on neurons in AD^59^. *AMIGO3* is among genes uniquely associated with the positive domain in the aINS. *AMIGO3* triggers the inhibition of oligodendrocyte precursor cell maturation, myelin production and neurite outgrowth^60^. Similar, we found *NPAS4* showing a unique positive association with the positive domain in the aINS. *NPAS4* encodes a transcription factor that regulates a number of downstream genes such as *BDNF, NARP* and *KCNA1*, which mediate diverse effects of synaptic modulation and experience-dependent memory formation^61^. Two other genes uniquely upregulated in association with the social domain in the aINS are *NME8* and *E2F8. NME8*, encoding TXNDC3 is involved in cytoskeletal function and axonal transport and identified as late-onset AD risk gene from GWAS and meta-analysis^62^. E2F8 is a member of E2F transcription factor family that regulate the transition from G1 to S phase, and aberrant activation of neuronal cell cycle has also been postulated as a mechanism of neuronal loss in AD^63^. Lastly, IL6, a major inflammatory marker, showed an increasing expression pattern in association with the negative domain in the aINS. This is consistent with previous meta-analyses showing positive association between inflammatory markers and depression^64^.

Several FDR-significant up- and downregulated genes were uniquely associated with individual clinical domains in the dACG. For example, *CNTF*, encoding a neurotrophic factor involved in neurotransmitter synthesis and neurite outgrowth, was uniquely upregulated in the social domain. *PINX1* was upregulated across the positive domain but downregulated across the cognitive and negative domain in the dACG. Interestingly it shows a unique upregulation with arousal in BA16 (*see above*). *SLC5A1*, encoding the sodium/glucose cotransporter SGLT1 and *LCN2*, encoding a glycoprotein expressed in reactive microglia and astrocytes in AD, were uniquely associated with the positive domain our cohort^65^. Genes associated with the negative domain and prior evidence for a role in AD included *IL1RL1, LIF*, and *FGB*^*66-68*^.

Although the number of overlapping DEGs across clinical domains was small (**Figure 2 C and D**), these genes could point to molecular mechanisms that influence a broader set of symptoms in AD. Common upregulated genes across all domains (including the cognitive domain) in the aINS indicate the overall activation of processes related to immune activation, which is an important mechanism contributing to AD pathogenesis and progression. For example, we found *CXCL10*, a chemokine that can mediate immune activation by binding to its receptor *CXCR3* and activate and recruit leukocytes^69^. We also found *MUC13*, which promotes NF-kB activity and leads to increased production of IL-8^70^. On the other hand, as a subtype of mucins, mature MUC13 can provide numerous glycosylation sites with its N-terminus located on the cell surface, and N-linked glycosylation has been recently reported to affect the progression of AD^71^. In total, we detect n=70 genes upregulated at p_FDR_<0.05 across at least two domains. In contrast, n=70 downregulated genes at p_FDR_<0.05 are shared across at least two domains (**Supplementary Data 6**).

In the dACG, several genes including *USP17L11, USP17L17, USP17L26*, and *USP17L28* were upregulated across different clinical domains. They belong to the deubiquitinating enzyme (DUB) family of genes and are involved in regulating the removal of ubiquitin molecules from proteins^72^. The ubiquitin-proteasome system (UPS) maintains mitochondrial homeostasis by regulating mitochondrial proteome and mitophagy^73^. UPS impairment and mitochondrial dysfunction have been implicated as hallmarks of aging and associated with neurodegenerative diseases such as Alzheimer’s disease and Parkinson disease^74,75^. Notably, *MT-RNR2*, encoding the polypeptide Humanin, is also upregulated across the cognitive, arousal, and social domain in the dACG^76^. In total, we detect n=66 genes upregulated at pFDR<0.05 across at least two domains. In contrast, n=78 downregulated genes at pFDR<0.05 are shared across at least two domains (**Supplementary Data 6**).

At the level of regulatory pathways, we found a more congruent profile with shared pathways related to the innate immune response defining upregulated gene profiles, and GTPase activity defining downregulated gene profiles across domains and (**Figure 2 E and F, Supplemental Data 7**). This contrasting result compared to the regulation of individual genes may reflect the overall strong impact of immune dysregulation, vesicle trafficking, and cell cycle regulation in AD progression.

In summary, the use of NLP-derived dimensional phenotypes may provide more specific insight into the underlying biology of AD. Each clinical domain score was associated with a distinct pattern of DEGs with a limited number of DEGs shared across domains. However, changes across clinical domains may be driven by shared functional pathways with a focus on immune system dysregulation.

## Acknowledgment statement

This work was supported by P50MH115874, R01MH120991, German Research Foundation #413501650, P50NM119467, Alzheimer’s Association #AACSFD-23-1149842, Eric Dorris Memorial Fellowship, Rappaport Mental Health Research Award, R01HD102974, and R01AG070704.

## Supplemental Figure Legends

**Figure S1. Identification of covariates in GLM differential expression model**

(**A-D**) Principal component analysis of group, brain region, sex, and sequence batch. Colors indicate sample subgroups under different conditions. (**E**) Pearson correlation matrix between the first five PCs from principal component analysis, potential variables of interest, and the first two surrogate variables from surrogate variable analysis. Colors indicate positive (red) and negative (blue) correlations. Dot size in the upper-right panel and numbers in the lower-left panel indicate the significant correlation coefficients.

## References

1 Sabates, J. et al. The Associations Between Neuropsychiatric Symptoms and Cognition in People with Dementia: A Systematic Review and Meta-Analysis. Neuropsychology Review 34, 581–597, doi:10.1007/s11065-023-09608-0 (2024).

2 Zhao, Q. F. et al. The prevalence of neuropsychiatric symptoms in Alzheimer’s disease: Systematic review and meta-analysis. J Affect Disord 190, 264–271, doi:10.1016/j.jad.2015.09.069 (2016).

3 Braak, H., Ra, d. V., En, J. S., Bratzke, H. & Braak, E. v. J. P. i. b. r. Neuropathological hallmarks of Alzheimer’s and Parkinson’s diseases. 117, 267–285 (1998).

4 Strooper, B. D. & Karran, E. J. C. The Cellular Phase of Alzheimer’s Disease. 164, 603–615 (2016).

5 Colangelo, V. et al. Gene expression profiling of 12633 genes in Alzheimer hippocampal CA1: transcription and neurotrophic factor down-regulation and up-regulation of apoptotic and pro-inflammatory signaling. 70, 462–473 (2002).

6 Zhang, B. et al. Integrated systems approach identifies genetic nodes and networks in late-onset Alzheimer’s disease. 153, 707–720 (2013).

7 Miller, J. A., Woltjer, R. L., Goodenbour, J. M., Horvath, S. & Geschwind, D. H. J. G. m. Genes and pathways underlying regional and cell type changes in Alzheimer’s disease. 5, 1–17 (2013).

8 Bossers, K. et al. Concerted changes in transcripts in the prefrontal cortex precede neuropathology in Alzheimer’s disease. 133, 3699–3723 (2010).

9 Neff, R. A. et al. Molecular subtyping of Alzheimer’s disease using RNA sequencing data reveals novel mechanisms and targets. 7 (2021).

10 Lam, B. et al. Clinical, imaging, and pathological heterogeneity of the Alzheimer’s disease syndrome. 5, 1–14 (2013).

11 Annese, A. et al. Whole transcriptome profiling of Late-Onset Alzheimer’s Disease patients provides insights into the molecular changes involved in the disease. 8, 4282 (2018).

12 Van Rooij, J. G. et al. Hippocampal transcriptome profiling combined with protein-protein interaction analysis elucidates Alzheimer’s disease pathways and genes. 74, 225–233 (2019).

13 Augustine, J. R. J. B. r. r. Circuitry and functional aspects of the insular lobe in primates including humans. 22, 229–244 (1996).

14 Showers, M. J. C. & Lauer, E. W. J. J. o. C. N. Somatovisceral motor patterns in the insula. 117, 107–115 (1961).

15 Kurth, F. et al. A link between the systems: functional differentiation and integration within the human insula revealed by meta-analysis. 214, 519–534 (2010).

16 Molnar-Szakacs, I., Uddin, L. Q. J. N. & Reviews, B. Anterior insula as a gatekeeper of executive control. 139, 104736 (2022).

17 Gogolla, N. J. C. B. The insular cortex. 27, R580–R586 (2017).

18 Dai, Z. et al. Identifying and mapping connectivity patterns of brain network hubs in Alzheimer’s disease. 25, 3723–3742 (2015).

19 Fathy, Y. Y. et al. Differential insular cortex sub-regional atrophy in neurodegenerative diseases: a systematic review and meta-analysis. 14, 2799–2816 (2020).

20 Jones, S. A. et al. Altered frontal and insular functional connectivity as pivotal mechanisms for apathy in Alzheimer’s disease. 119, 100–110 (2019).

21 Liu, X. et al. Altered functional connectivity of insular subregions in Alzheimer’s disease. 10, 107 (2018).

22 Gazestani, V. et al. Early Alzheimer’s disease pathology in human cortex involves transient cell states. 186, 4438-4453. e4423 (2023).

23 Mathys, H. et al. Single-cell atlas reveals correlates of high cognitive function, dementia, and resilience to Alzheimer’s disease pathology. 186, 4365-4385. e4327 (2023).

24 McCoy Jr, T. H. et al. High throughput phenotyping for dimensional psychopathology in electronic health records. 83, 997–1004 (2018).

25 Insel, T. et al. Vol. 167 748–751 (Am Psychiatric Assoc, 2010).

26 Cuthbert, B. N. J. W. p. The RDoC framework: facilitating transition from ICD/DSM to dimensional approaches that integrate neuroscience and psychopathology. 13, 28–35 (2014).

27 Carcone, D. & Ruocco, A. C. J. F. i. c. n. Six years of research on the national institute of mental health’s research domain criteria (RDoC) initiative: a systematic review. 11, 46 (2017).

28 Braak, H. & Braak, E. Staging of Alzheimer’s disease-related neurofibrillary changes. Neurobiol Aging 16, 271–278; discussion 278-284, doi:10.1016/0197-4580(95)00021-6 (1995).

29 Dobin, A. et al. STAR: ultrafast universal RNA-seq aligner. Bioinformatics 29, 15–21, doi:10.1093/bioinformatics/bts635 (2013).

30 Danecek, P. et al. Twelve years of SAMtools and BCFtools. GigaScience 10, giab008, doi:10.1093/gigascience/giab008 (2021).

31 Patro, R., Duggal, G., Love, M. I., Irizarry, R. A. & Kingsford, C. Salmon provides fast and bias-aware quantification of transcript expression. Nature Methods 14, 417–419, doi:10.1038/nmeth.4197 (2017).

32 Soneson, C., Love, M. I. & Robinson, M. D. Differential analyses for RNA-seq: transcript-level estimates improve gene-level inferences. F1000Res 4, 1521, doi:10.12688/f1000research.7563.2 (2015).

33 Love, M. I., Huber, W. & Anders, S. Moderated estimation of fold change and dispersion for RNA-seq data with DESeq2. Genome Biology 15, 550, doi:10.1186/s13059-014-0550-8 (2014).

34 Vogelgsang, J. S. et al. Dimensional clinical phenotyping using post-mortem brain donor medical records: Association with neuropathology. 2023.2005. 2004.539430 (2023).

35 Fischer, D. S., Theis, F. J. & Yosef, N. Impulse model-based differential expression analysis of time course sequencing data. Nucleic Acids Research 46, e119–e119, doi:10.1093/nar/gky675 (2018).

36 Leek, J. T., Johnson, W. E., Parker, H. S., Jaffe, A. E. & Storey, J. D. The sva package for removing batch effects and other unwanted variation in high-throughput experiments. Bioinformatics 28, 882–883, doi:10.1093/bioinformatics/bts034 (2012).

37 Zhou, Y. et al. Metascape provides a biologist-oriented resource for the analysis of systems-level datasets. Nature Communications 10, 1523, doi:10.1038/s41467-019-09234-6 (2019).

38 Vogelgsang, J. et al. Dimensional clinical phenotyping using post-mortem brain donor medical records: post-mortem RDoC profiling is associated with Alzheimer’s disease neuropathology. Alzheimer’s & Dementia: Diagnosis, Assessment & Disease Monitoring 15, e12464, 10.1002/dad2.12464 (2023).

39 Mufson, E. J. et al. Braak stage and trajectory of cognitive decline in noncognitively impaired elders. Neurobiology of Aging 43, 101–110, 10.1016/j.neurobiolaging.2016.03.003 (2016).

40 Nelson, P. T., Braak, H. & Markesbery, W. R. Neuropathology and cognitive impairment in Alzheimer disease: a complex but coherent relationship. J Neuropathol Exp Neurol 68, 1–14, doi:10.1097/NEN.0b013e3181919a48 (2009).

41 Schüller, M., Jenne, D. E. & Voltz, R. J. J. o. N. The human PNMA family: Novel neuronal proteins implicated in paraneoplastic neurological disease. 169, 172–176 (2005).

42 Pang, S. W., Lahiri, C., Poh, C. L. & Tan, K. O. J. C. s. PNMA family: Protein interaction network and cell signalling pathways implicated in cancer and apoptosis. 45, 54–62 (2018).

43 Xiong, J. et al. FSH blockade improves cognition in mice with Alzheimer’s disease. Nature 603, 470–476, doi:10.1038/s41586-022-04463-0 (2022).

44 Fisher, D. W., Bennett, D. A. & Dong, H. J. N. o. a. Sexual dimorphism in predisposition to Alzheimer’s disease. 70, 308–324 (2018).

45 Heneka, M. T. et al. Neuroinflammation in Alzheimer’s disease. 14, 388–405 (2015).

46 Eteleeb, A. M. et al. Brain high-throughput multi-omics data reveal molecular heterogeneity in Alzheimer’s disease. PLoS Biol 22, e3002607, doi:10.1371/journal.pbio.3002607 (2024).

47 Saura, C. A., Deprada, A., Capilla-López, M. D. & Parra-Damas, A. Revealing cell vulnerability in Alzheimer’s disease by single-cell transcriptomics. Seminars in Cell & Developmental Biology 139, 73–83, 10.1016/j.semcdb.2022.05.007 (2023).

48 Ross, C. A. & Margolis, R. L. Research Domain Criteria: Strengths, Weaknesses, and Potential Alternatives for Future Psychiatric Research. Mol Neuropsychiatry 5, 218–236, doi:10.1159/000501797 (2019).

49 Dalgleish, T., Black, M., Johnston, D. & Bevan, A. Transdiagnostic approaches to mental health problems: Current status and future directions. J Consult Clin Psychol 88, 179–195, doi:10.1037/ccp0000482 (2020).

50 Hengartner, M. P. & Lehmann, S. N. Why Psychiatric Research Must Abandon Traditional Diagnostic Classification and Adopt a Fully Dimensional Scope: Two Solutions to a Persistent Problem. Front Psychiatry 8, 101, doi:10.3389/fpsyt.2017.00101 (2017).

51 Fisher, D. W. et al. Unique transcriptional signatures correlate with behavioral and psychological symptom domains in Alzheimer’s disease. Translational Psychiatry 14, 178, doi:10.1038/s41398-024-02878-z (2024).

52 Pimplikar, S. W., Nixon, R. A., Robakis, N. K., Shen, J. & Tsai, L.-H. J. T. J. o. N. Amyloid-Independent Mechanisms in Alzheimer’s Disease Pathogenesis. 30, 14946–14954 (2010).

53 Dileep, V. et al. Neuronal DNA double-strand breaks lead to genome structural variations and 3D genome disruption in neurodegeneration. 186, 4404-4421.e4420 (2023).

54 Ferreira, A. C. et al. From the periphery to the brain: Lipocalin-2, a friend or foe? 131, 120–136 (2015).

55 Dekens, D. W. et al. Lipocalin 2 as a link between ageing, risk factor conditions and age-related brain diseases. 70, 101414 (2021).

56 Velasco, G. et al. Cloning and characterization of human MMP-23, a new matrix metalloproteinase predominantly expressed in reproductive tissues and lacking conserved domains in other family members. 274, 4570–4576 (1999).

57 Wang, X. et al. Distinct roles of telomerase activity in age-related chronic diseases: An update literature review. 167, 115553 (2023).

58 Chen, W.-D. et al. The circadian rhythm controls telomeres and telomerase activity. 451, 408–414 (2014).

59 Lyon, M. S. & Milligan, C. J. N. L. Extracellular heat shock proteins in neurodegenerative diseases: new perspectives. 711, 134462 (2019).

60 Foale, S., Berry, M., Logan, A., Fulton, D. & Ahmed, Z. J. N. r. r. LINGO-1 and AMIGO3, potential therapeutic targets for neurological and dysmyelinating disorders? 12, 1247 (2017).

61 Sun, X. & Lin, Y. J. T. i. n. Npas4: linking neuronal activity to memory. 39, 264–275 (2016).

62 Lambert, J.-C. et al. Meta-analysis of 74,046 individuals identifies 11 new susceptibility loci for Alzheimer’s disease. 45, 1452–1458 (2013).

63 Currais, A., Hortobágyi, T. & Soriano, S. J. A.The neuronal cell cycle as a mechanism of pathogenesis in Alzheimer’s disease. 1, 363 (2009).

64 Valkanova, V., Ebmeier, K. P. & Allan, C. L. J. J. o. a. d. CRP, IL-6 and depression: a systematic review and meta-analysis of longitudinal studies. 150, 736–744 (2013).

65 Ishida, N. et al. SGLT1 participates in the development of vascular cognitive impairment in a mouse model of small vessel disease. 727, 134929 (2020).

66 Jiang, Y. et al. An IL1RL1 genetic variant lowers soluble ST2 levels and the risk effects of APOE-ε4 in female patients with Alzheimer’s disease. 2, 616–634 (2022).

67 Soilu-Hänninen, M. et al. Expression of LIF and LIF receptor beta in Alzheimer’s and Parkinson’s diseases. 121, 44–50 (2010).

68 Bian, Z. et al. Accelerated accumulation of fibrinogen peptide chains with Aβ deposition in Alzheimer’s disease (AD) mice and human AD brains. 1767, 147569 (2021).

69 Michlmayr, D., McKimmie, C. S. J. I. j. o. i., cytokine & research, m. Role of CXCL10 in central nervous system inflammation. 1–18 (2014).

70 Sheng, Y. et al. MUC1 and MUC13 differentially regulate epithelial inflammation in response to inflammatory and infectious stimuli. 6, 557–568 (2013).

71 Zhao, J. & Lang, M. J. C. D. D. New insight into protein glycosylation in the development of Alzheimer’s disease. 9, 314 (2023).

72 Komander, D., Clague, M. J. & Urbé, S. J. N. r. M. c. b. Breaking the chains: structure and function of the deubiquitinases. 10, 550–563 (2009).

73 Ross, J. M., Olson, L. & Coppotelli, G. J. I. j. o. m. s. Mitochondrial and ubiquitin proteasome system dysfunction in ageing and disease: two sides of the same coin? 16, 19458–19476 (2015).

74 Hanpude, P., Bhattacharya, S., Dey, A. K. & Maiti, T. K. J. I. l. Deubiquitinating enzymes in cellular signaling and disease regulation. 67, 544–555 (2015).

75 Harrigan, J. A., Jacq, X., Martin, N. M. & Jackson, S. P. J. N. r. D. d. Deubiquitylating enzymes and drug discovery: emerging opportunities. 17, 57–78 (2018).

76 Niikura, T. J. B. e. B. A.-G. S. Humanin and Alzheimer’s disease: The beginning of a new field. 1866, 130024 (2022).

